# SVkhor: a unified framework for structural variant integration across long-read, short-read, and optical genome mapping data

**DOI:** 10.64898/2026.07.30.26359319

**Authors:** Edris Sharif Rahmani, Quentin Thomas, Emilie Tisserant, Valentin Vautrot, Anthony Auclair, Fridolin-Zinsou Hounnondaho, Estella Castillon, Laurence Faivre, Christel Thauvin-Robinet, Antonio Vitobello, Yannis Duffourd

## Abstract

**Summary:** Multi-technology human genome structural variant (SV) discovery is challenged by differences in breakpoint resolution, allele representation, SV annotation, and VCF structure across various callers and platforms. Here, we present SVkhor, a software framework designed to merge outputs from multiple callers within each technology and integrate SV callsets across available short-read sequencing, long-read sequencing, and optical genome mapping data. SVkhor addresses these challenges through caller-aware normalization, within-technology merging, and cross-technology integration, producing compact, source-annotated SV catalogs suitable for benchmarking and downstream interpretation. Benchmarking using HG002 and analysis of a clinical trio demonstrate that SVkhor reduces redundant caller-level complexity while preserving technology-specific evidence, enabling the transition from heterogeneous SV callsets to interpretable sample- and family-level SV catalogs.

**Availability and implementation:** SVkhor is implemented as a Linux command-line workflow. The source code, documentation, and example workflows are accessible at http://gitlab.gad-bioinfo.org/gad-public/svkhor under the MIT license.

**Supplementary information:** Supplementary data are accessible online.

## 1 Introduction

Structural variants (SVs) are genomic variants involving DNA segments of at least 50 base pairs in length, including deletions, insertions, duplications, inversions, translocations, and complex rearrangements. They represent a major source of human genetic variation, influencing genome function and contributing to human disease (Mahmoud et al. 2019, Collins et al. 2020). Accurate discovery and comparison of SVs are difficult because they depend on sequencing technology, genomic context, variant-calling methods, and the representation of the same biological events across callsets (Zook et al. 2020, Nardone et al. 2025).

Short-read (SR) sequencing remains widely used for genome analysis, but its ability to detect SV is limited by read length and varies across variant classes and genomic contexts, with reduced sensitivity in repetitive regions such as segmental duplications and simple repeats. Long-read (LR) sequencing improves SV detection and breakpoint resolution, whereas optical genome mapping (OGM), a non-sequencing-based technology, provides long-range structural information and is particularly suited to study large or complex rearrangements and for SV phasing (Zhao et al. 2021, De Clercq et al. 2024, Fu et al. 2026). Together, these technologies provide complementary data across different SV classes and genomic regions, motivating integrative approaches that combine heterogeneous SV callsets.

Despite this complementarity, the routine integration of LR, SR, and OGM callsets remains challenging. These technologies differ in read or molecule scale and breakpoint resolution, as well as the SV classes they detect (Zook et al. 2020, Nardone et al. 2025). In addition, variant callers and downstream SV analysis methods can represent related SVs differently and rely on different matching criteria, complicating comparison and integration across callsets (Tan et al. 2015, English et al. 2022, Nardone et al. 2025). Existing tools address parts of this challenge by clustering or merging related SV records. SURVIVOR and Jasmine primarily rely on positional similarity criteria for SV clustering, whereas SVanalyzer and PanPop incorporate sequence-level information to improve SV comparison and clustering (Jeffares et al. 2017, Zook et al. 2020, Kirsche et al. 2023, Zheng et al. 2024). However, integrating multi-technology LR, SR, and OGM callsets requires dedicated processing and source-aware interpretation.

We developed SVkhor to address this gap in SV analysis. It combines caller-aware VCF normalization, graph-based merging within each technology, and cross-technology integration of LR, SR, and OGM evidence. The output is a unified, source-annotated, sample-level SV catalog.

## 2 Methods

SVkhor merges SV calls from multiple callers within each technology, then integrates the resulting callsets across the available LR, SR, and OGM data for each sample. The workflow begins with caller-specific VCF files and proceeds through three main steps (Figure 1). First, svnorm standardizes heterogeneous VCF records by harmonizing SV types, genomic spans, allele descriptions, chromosome names, and source-call annotations. Second, for each technology, svmerge identifies equivalent normalized records and merges them into a technology-specific callset. Third, techmerge compares the technology-specific callsets and consolidates matching evidence into a single sample-level SV catalog annotated with its supporting technologies.

**Figure 1.**
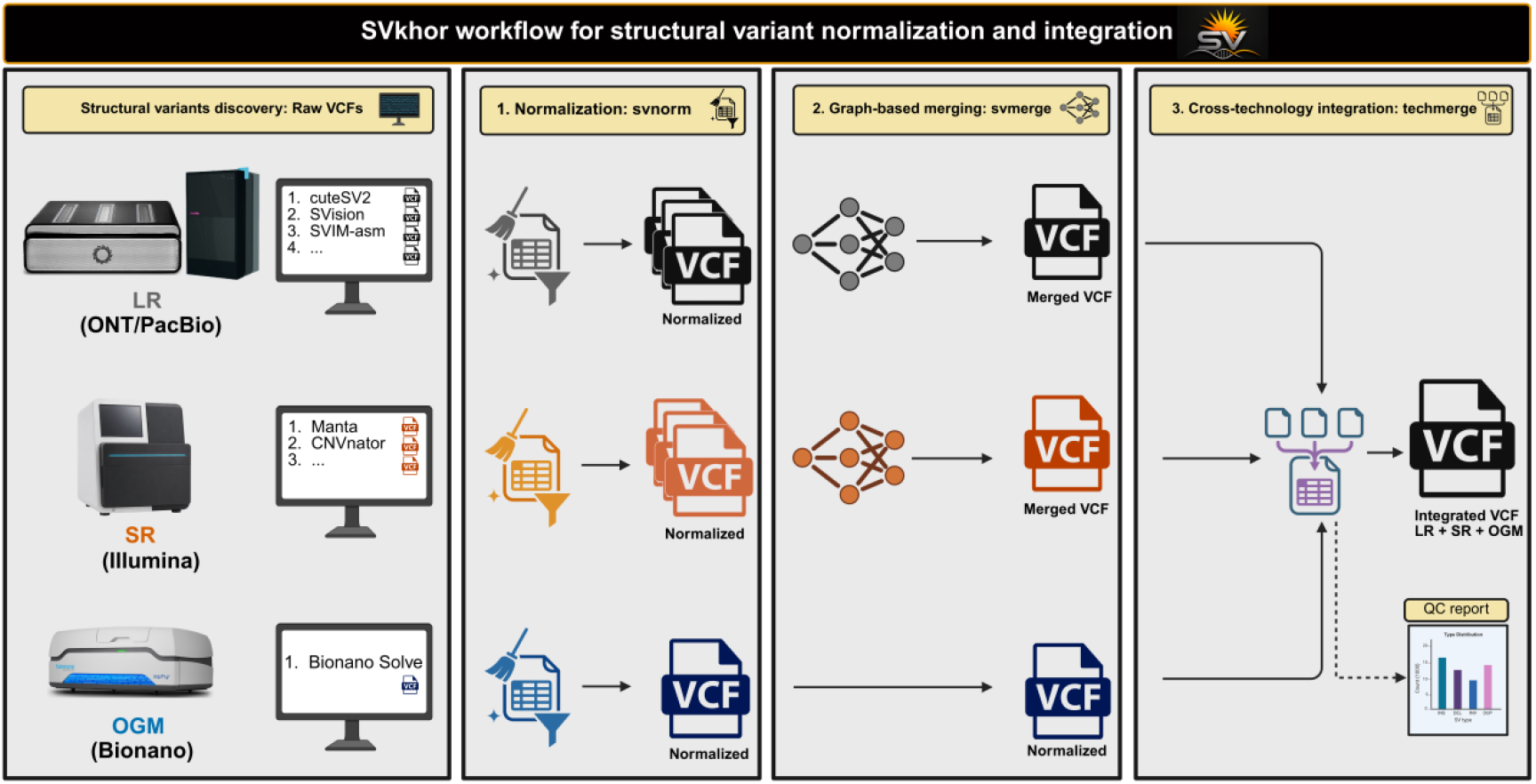
Overview of the SVkhor workflow. Caller-specific VCF files from LR sequencing, SR sequencing, and OGM are initially standardized by svnorm. Subsequently, normalized LR and SR calls are merged within each technology by svmerge. Finally, techmerge integrates LR, SR, and OGM evidence into a single source-annotated VCF. An optional quality-control report summarizes technology support and integration outcomes.

### 2.1 VCF normalization

Svnorm standardizes caller-specific VCF records prior to merging. For each variant, the SV type is extracted from the INFO field when available. If unavailable, the SV type is inferred from the symbolic ALT allele or from the REF/ALT length difference for sequence-resolved insertions and deletions. Genomic span and SV length are derived from END and SVLEN when present, or calculated from allele lengths when possible. Records with ambiguous or contradictory type, position, span, or length are not carried forward into the normalized callset because svmerge requires consistent SV type and genomic span for event matching; such records can be written to a rejection file for review. Chromosome names are converted to the corresponding contig names in the reference FASTA index. Normalized records are written in reference-contig and genomic coordinate order. Multiallelic records are divided into one alternate allele per record, with genotype fields remapped to the retained allele. For literal sequence-resolved deletion/insertion (DEL/INS) records, svnorm applies reference-aware allele normalization according to the left-alignment and parsimony principles of VCF normalization (Tan et al. 2015): shared suffixes and prefixes are trimmed, alleles are left-extended with the preceding reference base when required to keep both alleles non-empty, and POS, END, and SVLEN are recalculated from the normalized representation.

### 2.2 Within-technology merging

Svmerge clusters equivalent normalized records separately within each technology. Each SV is compared only with records on the same chromosome and of the same type. Candidate matches are evaluated using SV-type-specific criteria, including breakpoint and interval agreement for span-defined events, Edlib-based sequence similarity when resolved alleles are available (Šošić and Šikić 2017), orientation agreement for breakend-like events, and copy-number compatibility for copy number variants (CNVs). Svmerge can adjust the matching stringency in low-mappability and segmental-duplication regions, where breakpoint positions are less reliably resolved (Ahsan et al. 2023). Compatible records are connected in an SV graph, and each connected component is represented by a single merged VCF record.

### 2.3 Cross-technology integration

Techmerge integrates the LR, SR, and OGM callsets into a single per-sample SV catalog. LR calls serve as the backbone for cross-technology integration because they typically provide higher-resolution SV records, particularly for breakpoint placement and allele representation. SR calls are matched to LR events using breakpoint proximity, SV-length similarity, and allele-sequence similarity when explicit alleles are available. Unmatched SR calls are retained as SR-specific events after filtering weak SR-only records that fall within the local neighborhood of stronger LR-supported events. OGM calls are then matched with nearby LR or SR events within the same genomic interval, allowing multiple compatible read-based events to collectively explain an OGM signal. Concordant OGM calls are added as supporting evidence, while unmatched or discordant calls are retained as OGM-specific events. Final records retain explicit technology-support annotations, indicating whether each event is supported by LR, SR, OGM, or a combination of these evidence sources.

### 2.4 Dataset and preprocessing

SVkhor was evaluated using the Genome in a Bottle (GIAB) HG002 sample, incorporating Illumina short-read sequencing, Oxford Nanopore long-read sequencing, and Bionano optical genome mapping data (Zook et al. 2020). All sequencing datasets were aligned to a GRCh38 reference that included decoy contigs and had pseudoautosomal regions on chromosome Y masked to minimize ambiguous mapping between homologous X and Y regions. Illumina reads were processed with fastp v1.3.2 (Chen 2025) and aligned using BWA-MEM (Li 2013). Oxford Nanopore reads were filtered to retain those of at least 300 base pairs, aligned with minimap2 v2.30 (Li 2018), and additionally corrected with Ratatosk v0.9.0 (Holley et al. 2021) using the matched SR data. Corrected long reads were also assembled *de novo* with Flye v2.9.6 (Kolmogorov et al. 2019), and the resulting contigs were aligned using minimap2 with the asm5 preset. Alignment and coverage statistics were obtained using samtools flagstat and samtools coverage v1.22.1 (Li et al. 2009).

### 2.5 Structural variant discovery

Structural variant discovery was performed separately for each data type. For SR data, CNVnator v0.4.1 (Abyzov et al. 2011) and Manta v1.6.0 (Chen et al. 2016) were used. For LR data, SV calls were generated using cuteSV2 v2.1.1 (Jiang et al. 2020), Dysgu v1.8.7 (Cleal and Baird 2022), SVIM-asm v1.0.3 (Heller and Vingron 2020), and SVision v1.4 (Lin et al. 2022), while variable number tandem repeats (VNTRs) were identified using LongTR v1.2 (Ziaei Jam et al. 2024). OGM SVs and CNVs were generated using Bionano Solve v3.8.2.1 with the *de novo* assembly pipeline, including auto-noise rescaling and alignment to the hg38C_BSPQI *in silico* reference map (Zook et al. 2016, Bionano Genomics 2024). All callers were run with default parameters, except that SV discovery or downstream caller-level filtering was restricted to variants of at least 50 base pairs.

### 2.6 Benchmarking and tool comparison

Benchmarking was limited to deletions and insertions of at least 50 base pairs, consistent with the scope of the GIAB HG002 (v5.0q) benchmark, which contains 28,123 high-confidence events highlighted in the BED file. Merged callsets were evaluated using Truvari v5.4.0 with --sizemin 50, --pctsize 0.5, --pctseq 0.5, and -- refdist 1000 (English et al. 2022). SVkhor was compared with a no-merge baseline generated by concatenating normalized VCF files using bcftools concat (Danecek et al. 2021), without applying SV equivalence matching or clustering, and with existing SV merging tools, including Jasmine v1.1.5 (Kirsche et al. 2023), SURVIVOR v1.0.7 (Jeffares et al. 2017), SVanalyzer v0.36 (Zook et al. 2020), and PanPop (Zheng et al. 2024). Comparisons were performed under three input conditions: LR-only, SR-only, and integrated LR+SR+OGM. Truvari genotype (GT) concordance was reported only for VCFs with informative GT values; site-only or missing-GT outputs were marked as not applicable.

### 2.7 Clinical use case

To illustrate the application of SVkhor in a diagnostic setting, we applied the pipeline to a neurodevelopmental-disorder trio from our laboratory for which LR, SR, and OGM data were available. The case was processed in accordance with sections 2.4 and 2.5. SVkhor was applied using the same normalization, within-technology merging, and cross-technology integration strategy described above. Following generation of sample-level integrated SV catalogs, the svmerge cohort-integration mode was used to combine the three family members into a non-redundant family-level SV catalog.

## 3 Results

### 3.1 HG002 multi-technology inputs

The HG002 dataset provided high-coverage SR, LR, and OGM evidence for SV integration. After filtering, the Illumina dataset retained 5.89 billion reads, achieving a 99.75% mapping rate and a depth of 267×. The Oxford Nanopore dataset comprised 156.5 GB of sequence data, with a read N50 of 48.4 kb, and reached a depth of 41.3× following alignment. Ratatosk correction improved the mean base quality from Q30.9 to Q35.3, while maintaining comparable mapping quality and coverage. For OGM, 1.28 million Bionano molecules were retained after auto-noise rescaling, of which 780,185 aligned to the reference map, yielding 63.56× effective reference coverage (Supplementary Table 1).

Structural variant discovery produced heterogeneous caller outputs across technologies, ranging from 4,567 Bionano records to 197,702 Manta records. After preprocessing for deletions and insertions of at least 50 base pairs using svnorm, the standardized SVkhor input included 47,567 SR records, 135,779 LR records, and 4,229 OGM records (Supplementary Table 2). Records were rejected primarily due to being outside the benchmark scope, such as unsupported SV classes, noncanonical contigs, variants below 50 base pairs, ambiguous breakend or partner-contig representations, or sequence-resolved records with no effective SV length change (Supplementary Table 3).

### 3.2 HG002 benchmarking and tool comparison

Using these standardized inputs, SVkhor generated merged callsets for both single- and multi-technology analyses. Within-technology svmerge generated 76,650 LR and 41,254 SR records, with 60,320 LR and 23,005 SR records retained after benchmark filtering. LR merging achieved the highest recall among the dedicated single-technology merging methods, with 0.923 recall and an F1 score of 0.587, whereas SR-only merging recovered fewer benchmark events, consistent with the lower performance observed across all tools in the SR-only setting.

Cross-technology integration with techmerge generated 110,941 HG002 DEL/INS records, with 17.2% supported by more than one technology (Supplementary Figure 1). The Truvari-evaluated LR+SR+OGM set identified 26,151 true positives, resulting in a recall of 0.930 and an F1 score of 0.499. Compared with naive bcftools concatenation, SVkhor recovered 98.6% of true positives while reducing false positives by 58.2% (Figure 2A and Supplementary Figure 2).

**Figure 2.**
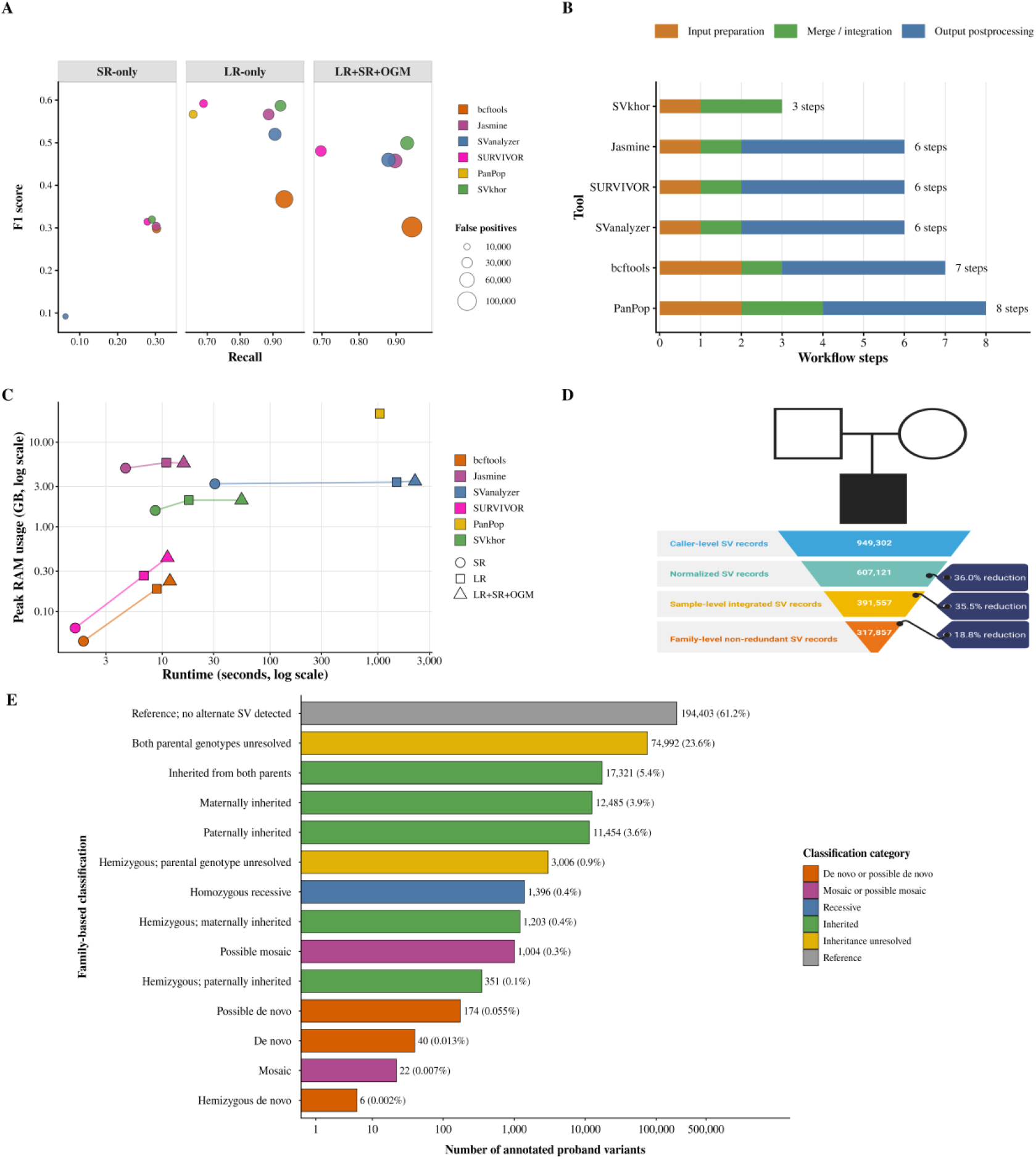
Benchmarking, workflow complexity, computational performance, and clinical application of SVkhor. (A) Truvari benchmarking of SR-only, LR-only, and integrated LR+SR+OGM callsets against the GIAB HG002 GRCh38 DEL/INS benchmark. Each point represents a tool within a benchmarking scenario; the x-axis indicates recall, the y-axis indicates F1 score, and point size reflects the number of false positives. Higher values on the x- and y-axes correspond to greater recall and F1 score, respectively, while larger points indicate a higher false-positive burden. Missing points denote cases where no benchmarkable output was produced or the metric was undefined. (B) Number and type of workflow steps required to generate benchmark-ready VCF files for SVkhor and other evaluated tools, grouped into input preparation, merging or integration, and output postprocessing. (C) Runtime and peak memory usage for SR-only, LR-only, and LR+SR+OGM analyses, with both axes presented on a logarithmic scale. (D) Application of SVkhor to a neurodevelopmental-disorder trio, demonstrating reduction from caller-level SV records to normalized records, sample-level integrated SV records, and a non-redundant family-level SV catalog. (E) Proband-centered inheritance classification of 317,857 non-redundant trio-wide SV events. Bars show event counts, with percentages calculated relative to the complete family-level catalog; colors indicate broader classification groups. The “Both parental genotypes unresolved” category denotes autosomal or other diploid events for which both parental GTs were unresolved. Inherited from both parents indicates an alternate SV genotype in the proband and both parents. Hemizygous parental genotype unresolved denotes a male chrX or chrY event with an unresolved genotype in the relevant transmitting parent. Mosaic indicates a low-variant-allele-fraction proband event with callable reference parents, whereas possible mosaic indicates the same proband signal with incomplete parental genotype information. Possible *de novo* denotes an alternate proband genotype with one reference parent and one unresolved parent.

Jasmine and SVanalyzer maintained recall in both LR-only and LR+SR+OGM settings; however, SVkhor achieved higher sensitivity and superior overall precision–recall balance. SURVIVOR produced fewer false positives and lower sensitivity; its LR-only F1 score is reported in Figure 2A. PanPop was evaluated only on sequence-resolved records, consistent with its workflow design. Although 65.7% of SR+LR+OGM raw records met these criteria, the combined analysis did not yield an output suitable for Truvari benchmarking, reflecting the heterogeneous structure of the input data (Figure 2A and Supplementary Figure 2). Among all genotype-evaluable tools, SVkhor achieved the highest genotype concordance in both LR-only and integrated LR+SR+OGM analyses, with values of 0.776 and 0.777, respectively (Supplementary Figure 2).

### 3.3 Workflow complexity and computational performance

SVkhor required three workflow steps to generate a benchmark-ready merged VCF: svnorm, svmerge, and techmerge, with outputs produced as bgzipped VCF files that are sorted and indexed. In contrast, the external tools required six to eight workflow steps, including sequence-resolved filtering for PanPop, VCF header or sample-column repair as needed, multiallelic decomposition, sorting, compression, and indexing (Figure 2B and Supplementary Table 4).

Runtime and memory profiling was performed using up to four threads, where supported. SVkhor was not the fastest method in terms of absolute wall-clock time, but it was substantially faster than the most computationally intensive evaluated tools and required less memory than Jasmine, SVanalyzer, and PanPop. In the integrated LR+SR+OGM analysis, SVkhor completed the full workflow in 54.4 seconds with approximately 2.1 GB peak memory, making it about 41 times faster than SVanalyzer and about 19 times faster than the completed LR-only PanPop workflow. SVanalyzer and PanPop required approximately 1.7- and 10.5-fold more peak memory, respectively (Figure 2C).

### 3.4 Clinical trio SV integration with SVkhor

SVkhor processed 949,302 caller-level SV records across the three members of the neurodevelopmental-disorder trio and retained 607,121 normalized records after caller-aware harmonization. Cross-technology integration reduced these to 391,557 integrated SV records across the three individuals, and subsequent joint family integration produced 317,857 non-redundant trio-wide events (Figure 2D; Supplementary Tables 5 and 6). This unified family-level catalog enabled interpretation of inheritance patterns. We assigned the 317,857 events to 14 proband-centered classes (Figure 2E). Most events were classified as reference in the proband (61.2%) or had unresolved parental origin (23.6%), whereas 12.9% were maternally, paternally, or biparentally inherited. Recessive, mosaic, and *de novo* candidate classes together accounted for less than 1% of the catalog. This inheritance-aware classification supports downstream filtering and variant prioritization (Figure 2E).

## 4 Discussion

SVkhor was developed to convert heterogeneous outputs from LR, SR, and OGM callers into an interpretable, source-annotated SV catalog. SVkhor reduces redundant caller-specific records while retaining technology-support annotations, which addresses a recognized challenge in SV comparison when equivalent events are represented differently across technologies and callers (Zook et al. 2020, English et al. 2022, Nardone et al. 2025). This design preserves the source of each call while combining records that represent the same event.

In HG002, SVkhor reduced the false-positive burden while retaining most benchmark-supported events. In comparison, the no-merge bcftools concat baseline preserved slightly more true positives but also retained numerous benchmark-unmatched or redundant records due to the absence of SV equivalence matching. The observed stronger performance for LR-only data and weaker performance for SR-only data align with the higher breakpoint resolution of long-read sequencing and the established limitations of short-read SV detection in repetitive or complex genomic regions (Zhao et al. 2021, De Clercq et al. 2024).

Comparison with external tools indicates that performance is highly dependent on the underlying assumptions used during matching. Proximity-based tools are effective when breakpoint distance is an adequate criterion, while sequence-aware tools are more appropriate for resolved allele sequences and population-level merging (Kirsche et al. 2023, Zheng et al. 2024). However, integrating LR, SR, and OGM data must also address symbolic records and interval-based OGM evidence, underscoring the need for a framework that preserves heterogeneous evidence.

In addition to benchmark accuracy, SVkhor minimizes the manual preprocessing required to convert caller outputs into interpretable VCF files. Format repair, decomposition, sorting, compression, and indexing can introduce workflow differences between analyses. Consequently, a compact and reproducible workflow is advantageous for repeated sample analysis, where output consistency is as critical as runtime.

Analysis of the neurodevelopmental-disorder trio illustrates the utility of source-aware integration beyond benchmark datasets. For diagnostic review, the catalog should reduce redundant calls and allow joint inspection of genotype status and segregation across family members. This is particularly relevant for clinical SV interpretation, where LR sequencing and OGM offer complementary evidence for breakpoint resolution, phasing, and the interpretation of large or complex rearrangements (De Clercq et al. 2024, Fu et al. 2026).

This study was limited by the current scope of available benchmark resources. The GIAB HG002 high-confidence benchmark supports evaluation of deletions and insertions, but comparable truth sets are not yet available for all SV classes handled by SVkhor. Improved multi-class SV benchmarks would allow more complete assessment of tools designed for multi-technology SV integration. Future work should therefore extend evaluation to additional SV classes and independent benchmark samples. In summary, SVkhor provides a source-aware path from technology-specific caller outputs to sample- and family-level SV catalogs, with higher benchmark performance and fewer workflow steps.

## Data Availability

SVkhor source code, documentation, and example workflows are available at http://gitlab.gad-bioinfo.org/gad-public/svkhor. Public HG002 sequencing data and benchmark resources are available through the Genome in a Bottle consortium. Summary results for the clinical trio are provided in the manuscript and supplementary material. Individual-level clinical data are not publicly available because of participant confidentiality and institutional data-protection requirements.

https://www.nist.gov/programs-projects/genome-bottle

