## Supplementary material for "SVkhor: a unified framework for structural variant integration across long-read, short-read, and optical genome mapping data"

Supplementary Table 1. Summary of sequencing yield, read filtering, alignment, correction, assembly, and coverage metrics for the Illumina short-read, Oxford Nanopore long-read, and Bionano optical genome mapping datasets analyzed in this study. Reported values include sequencing output, filtering losses, alignment performance, breadth and depth of coverage, base and mapping quality summaries, molecule-level OGM statistics, the effect of hybrid correction on long-read data quality, and assembly contiguity metrics.

| Platform | Category | Metric | Value |
|---|---|---|---|
| Illumina short-read | Pre-filtering | Total reads | 6,270,610,276 |
| | | Total bases | 928.1 Gb |
| | | Bases at Q30 | 90.0% |
| | Post-filtering | Retained reads | 5,878,906,334 |
| | | Retained bases | 867.7 Gb |
| | | Bases at Q30 | 93.1% |
| | | Bases at Q20 | 98.1% |
| | Read filtering | Removed for low quality | 374,218,346 |
| | | Removed for short length | 15,930,410 |
| | | Removed for excessive Ns | 1,555,186 |
| | Alignment | Mapped reads | 5,891,317,767 |
| | | Mapping rate | 99.75% |
| | | Properly paired reads | 97.77% |
| | | Duplicate reads | 206,639,742 |
| | Coverage/quality | Weighted mean breadth | 95.0% |
| | | Weighted mean depth | 267× |
| | | Mean base quality | 35.9 |
| | | Mean mapping quality | 55.3 |
| Oxford Nanopore | Raw read set | Total reads | 19,328,993 |
| | | Total bases | 156.5 Gb |
| | | Read N50 | 48.4 kb |
| | | Read N75 | 19.6 kb |
| | Pre-alignment filtering | Minimum retained read length | 300 bp |
| | Alignment | Mapped reads | 7,674,987 |
| | | Mapping rate | 100% |
| | | Primary alignments | 7,019,139 |
| | | Supplementary alignments | 655,848 |
| | | Secondary alignments | 395,659 |
| | Coverage/quality | Weighted mean breadth | 94.4% |
| | | Weighted mean depth | 41.3× |
| | | Mean base quality | 30.9 |
| | | Mean mapping quality | 57.1 |
| Oxford Nanopore + Ratatosk | Alignment | Mapped reads | 7,635,683 |
| | | Primary alignments | 6,779,930 |
| | Coverage/quality | Weighted mean breadth | 93.5% |
| | | Weighted mean depth | 42.7× |
| | | Mean base quality | 35.3 |
| | | Mean mapping quality | 58.5 |
| | Assembly | Total contigs | 1,160 |
| | | Total assembly size | 2.86 Gb |
| | | Longest contig | 111.4 Mb |
| | Assembly contiguity | Contig N50 | 44.6 Mb |
| | | Contig N90 | 11.3 Mb |
| | Assembly Coverage/quality | Length-weighted mean contig coverage | 38.3× |
| Bionano OGM | Molecule statistics | Total molecules | 2,601,635 |
| | | Total molecule length | 507,197 Mbp |
| | | Average molecule length | 195 kb |

| | | | |
|---|---|---|---|
| | | Molecule N50 | 203 kb |
| | | Label density | 8.95 per 100 kb |
| | After auto-noise rescaling | Retained molecules | 1,284,682 |
| | | Retained molecule length | 339,248 Mbp |
| | | Average molecule length | 264 kb |
| | | Molecule N50 | 267 kb |
| | | Raw reference coverage | 109.85× |
| | Alignment to reference | Aligned molecules | 780,185 |
| | | Fraction aligned molecules | 0.607 |
| | | Total aligned molecule length | 195,340.6 Mbp |
| | | Effective reference coverage | 63.56× |
| | | Average aligned length | 250.4 kb |
| | | Average alignment confidence | 23.1 |

Notes: Ratatosk correction improved read-level base quality while leaving overall mapping and coverage metrics broadly similar. For long-read data, N50 is the read length such that 50% of all sequenced bases are contained in reads of that length or longer, and N75 is the corresponding read length threshold for 75% of sequenced bases.

Supplementary Table 2. Input callset sizes and INS/DEL-focused preprocessing counts. Raw VCF record counts are reported for each caller, along with the number of records retained after SVkhor svnorm normalization for INS/DEL variants of at least 50 base pairs and the corresponding bcftools-preprocessed counts used for comparator tools. Comparator inputs were preprocessed to restrict records to the DEL/INS ≥50 bp benchmark scope and to reduce caller-specific VCF-format differences where required. Final benchmark VCFs were then made Truvari-compatible by applying tool-specific postprocessing steps, including multiallelic decomposition, sorting, compression, indexing, and record-ID repair where needed. LongTR underwent caller-specific post-calling filtering before downstream analysis, retaining records with FMT/Q[0] >= 0.80, FMT/DP[0] >= 10, FMT/DFLANKINDEL[0] == 0, and ABS(INFO/BPDIFFS) >= 50. The retained LongTR records were subsequently used for INS/DEL-focused normalization and comparator preprocessing. For other sequencing callers, records were restricted to SVTYPE=INS/DEL with |SVLEN| >= 50. SVIM-asm calls were generated from de novo-assembled long-read contigs aligned to GRCh38 using minimap2, representing assembly-based structural-variant calls. For Bionano/Solve, the svnorm count reflects the harmonized INS/DEL-focused optical genome-mapping call set derived from the Solve export VCF. For LongTR, the svnorm count was higher than the raw VCF record count because multiallelic records were decomposed into biallelic records during normalization, resulting in 17,751 retained records.

| Technology | Caller | Raw VCF records | svnorm | bcftools filter |
|---|---|---|---|---|
| SR | CNVnator | 27,993 | 24,690 | 24,690 |
| | Manta | 197,702 | 22,877 | 23,100 |
| LR | cuteSV2 | 27,462 | 26,826 | 26,983 |
| | Dysgu | 45,166 | 43,031 | 44,136 |
| | LongTR | 14,692 | 17,751 | 14,692 |
| | SVIM-asm | 20,895 | 17,106 | 17,111 |
| | SVision | 31,373 | 31,065 | 31,075 |
| OGM | Bionano / Solve | 4,567 | 4,229 | 4,229 |

Supplementary Table 3. svnorm rejection categories for HG002 caller VCFs. The table summarizes the primary reasons for record exclusion during INS/DEL-focused svnorm preprocessing, applying a minimum structural variant (SV) length threshold of 50 bp. Counts are obtained from svnorm verbose logs and represent records rejected prior to generating the normalized DEL/INS callsets reported in Supplementary Table 2. For variant callers producing multiallelic records, rejection counts are provided after allele decomposition; thus, LongTR rejection counts correspond to normalized candidate alleles rather than original VCF rows.

| Technology | Caller | Rejected by svnorm | Primary rejection categories |
|---|---|---|---|
| SR | CNVnator | 3,303 | Unsupported SV type: 3,303 |
| | Manta | 174,825 | Size below 50 bp: 134,439; unsupported SV type: 34,401; noncanonical contig: 3,687; unresolved partner/breakend representation: 2,298 |
| LR | cuteSV2 | 636 | Unsupported SV type: 320; noncanonical contig: 316 |
| | Dysgu | 2,135 | Size below 50 bp: 1,103; unsupported SV type: 415; noncanonical contig: 364; unresolved partner/breakend representation: 253 |
| | LongTR | 4,344 | Size below 50 bp: 3,465; no effective SV length change: 879 |
| | SVIM-asm | 3,789 | Size below 50 bp: 3,655; unsupported SV type: 112; noncanonical contig: 20; unresolved partner/breakend representation: 2 |
| | SVision | 308 | Unsupported or compound SV type: 275; size below 50 bp: 23; noncanonical contig: 10 |
| OGM | Bionano / Solve | 338 | Unsupported SV type: 338; malformed breakends: 94 |

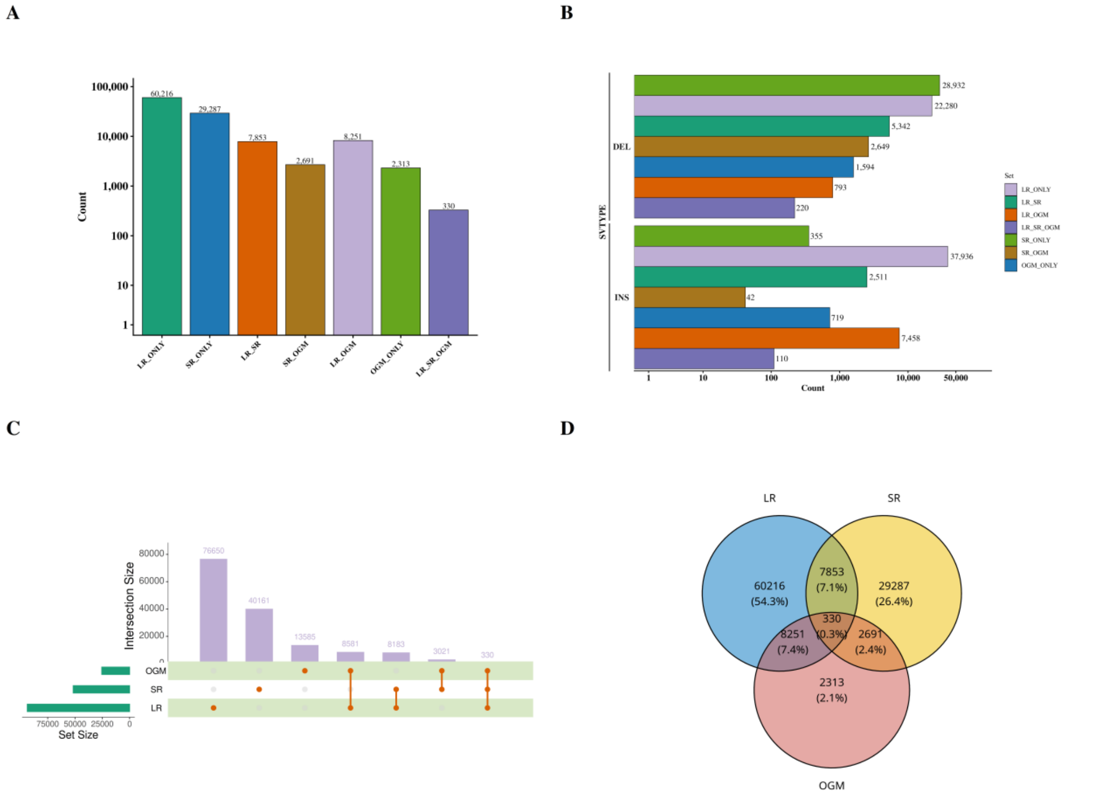


Supplementary Figure 1. Composition of the HG002 integrated insertion and deletion callset following integration of long-read (LR), short-read (SR), and optical genome mapping (OGM) data. The final integrated callset comprised 110,941 deletion/insertion (DEL/INS) variants. (A) Mutually exclusive source classes were primarily LR_ONLY (60,216; 54.3%) and SR_ONLY (29,287; 26.4%), followed by LR_OGM (8,251; 7.4%) and LR_SR (7,853; 7.1%). OGM-associated categories included LR_OGM (8,251; 7.4%), SR_OGM (2,691; 2.4%), OGM_ONLY (2,313; 2.1%), and LR_SR_OGM (330; 0.3%). (B) Deletions were slightly more prevalent than insertions in the integrated callset (61,810; 55.7% versus 49,131; 44.3%). LR_ONLY calls were enriched for insertions, whereas SR_ONLY and SR_OGM calls were predominantly deletions. (C) The UpSet-style summary presents inclusive modality support and overlaps: 76,650 calls had LR support, 40,161 had SR support, and 13,585 had OGM support. Inclusive overlaps included 8,183 LR+SR-supported calls, 8,581 LR+OGM-supported calls, 3,021 SR+OGM-supported calls, and 330 calls supported by all three technologies. (D) The Venn diagram shows the mutually exclusive partitioning of the integrated callset. Values and percentages in each Venn region are calculated relative to the complete integrated callset (n = 110,941), not relative to the total number of calls within each individual modality. Single-modality regions accounted for 91,816 records (82.8%), including LR_ONLY, SR_ONLY, and OGM_ONLY calls, whereas multi-modality regions accounted for 19,125 records (17.2%), including LR_SR, LR_OGM, SR_OGM, and LR_SR_OGM calls.

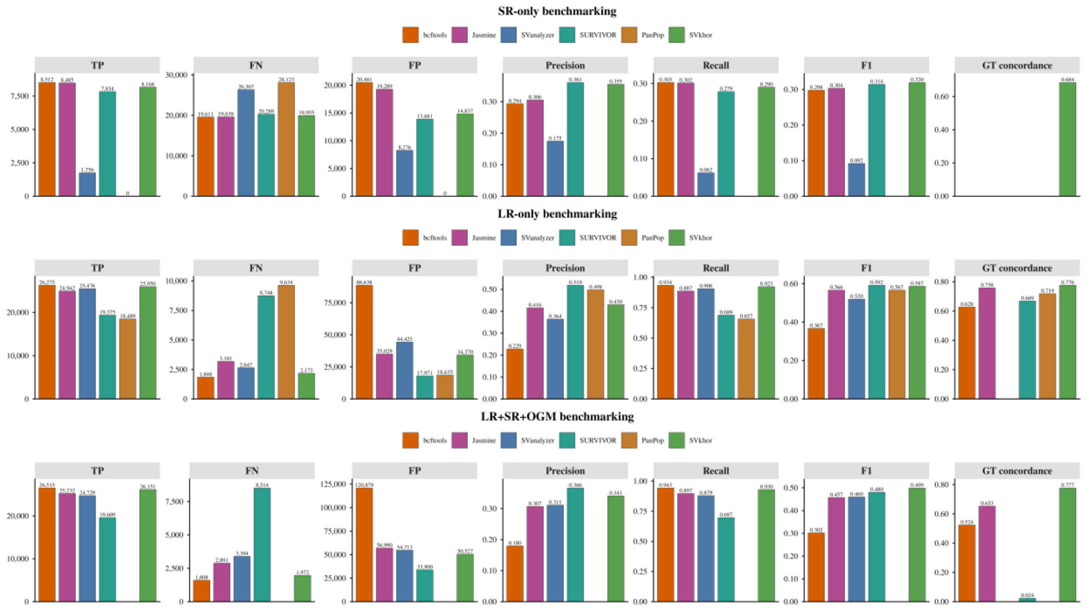


Supplementary Figure 2. Benchmarking comparison of SR-only, LR-only, and LR+SR+OGM callsets against the GIAB HG002 DEL/INS benchmark using Truvari. Bar plots show true positives (TP), false negatives (FN), false positives (FP), precision, recall, F1 score, and genotype (GT) concordance for bcftools, Jasmine, SVanalyzer, SURVIVOR, PanPop, and SVkhor. bcftools was included as a union-based non-merging baseline, whereas the other tools were evaluated as SV-merging or integration methods. In the SR-only setting, SVkhor achieved the highest F1 score among the dedicated merging methods and was the only method with evaluable GT concordance. In the LR-only setting, SVkhor achieved high recall and competitive F1, while SURVIVOR showed the lowest FP count and highest F1 among the dedicated merging tools. In the LR+SR+OGM setting, SVkhor provided the best overall balance among the dedicated integration methods, achieving the highest F1 score with high recall, whereas bcftools achieved the highest recall but produced substantially more FPs, consistent with its union-based strategy. PanPop produced a usable LR-only benchmark output, an empty SR-only benchmark VCF, and no benchmarkable LR+SR+OGM output. Zero-height bars indicate completed benchmarks with a true value of zero, whereas blank entries indicate metrics that were not evaluable because no benchmarkable output was produced or because the metric was undefined. GT concordance was plotted only for outputs with evaluable genotype information; site-only outputs, outputs lacking GT fields, and outputs with uninformative GT values were treated as not evaluable. Among genotype-evaluable outputs, SVkhor achieved the highest GT concordance in the SR-only, LR-only, and LR+SR+OGM comparisons.

Supplementary Table 4. Workflow steps required to generate benchmark-ready merged VCFs for SVkhor and comparator tools. This table summarizes the input preparation, merging or integration, and post-merge processing steps necessary prior to Truvari benchmarking. DEL/INS ≥50 bp filtering was applied to all comparator inputs, while svnorm incorporates this filtering during SVkhor caller-aware normalization. Additional post-merge steps, including VCF header or structure repair, multiallelic splitting, sorting, compression, and indexing, were applied only when required to produce a valid benchmark-ready VCF.

| Tool | Input preparation before merge | Merge / integration step | Post-merge processing before Truvari | Total major workflow steps |
|---|---|---|---|---|
| SVkhor | 1. Caller-aware normalization and benchmark-scope filtering with svnorm | 2. Within-technology merging with svmerge; 3. cross-technology integration with techmerge | Not required | 3 |
| bcftools | 1. Filter caller VCFs for the benchmark scope; 2. for LR+SR+OGM only, add missing HG002=./. sample fields to site-only SR records for concat compatibility | 3. Concatenate caller VCFs with bcftools concat | 4. Repair header definitions; 5. split multiallelic records; 6. sort; 7. bgzip and tabix index | 7 |
| SVanalyzer | 1. Filter caller VCFs for the benchmark scope | 2. Merge with SVanalyzer SVmerge | 3. Repair/rebuild VCF header definitions; 4. split multiallelic records; 5. sort; 6. bgzip and tabix index | 6 |
| PanPop | 1. Filter caller VCFs for the benchmark scope; 2. retain sequence-resolved records only | 3. Run PanPop PART pass 1; 4. run PanPop PART pass 2 | 5. Repair header definitions; 6. split multiallelic records; 7. sort; 8. bgzip and tabix index. | 8 |
| SURVIVOR | 1. Filter caller VCFs for the benchmark scope | 2. Merge with SURVIVOR | 3. Repair/rebuild VCF header definitions; 4. split multiallelic records; 5. sort; 6. bgzip and tabix index | 6 |
| Jasmine | 1. Filter caller VCFs for the benchmark scope | 2. Merge with Jasmine | 3. Repair/rebuild VCF header and body structure where required; 4. split multiallelic records; 5. sort; 6. bgzip and tabix index | 6 |

Supplementary Table 5. Caller-level SV counts for the clinical trio. For each family member, sequencing technology, SV caller, number of raw records loaded by svnorm, number of normalized records retained by SVkhor, and the distribution of normalized records by SV class are reported. Records classified as BND, CSV, DEL, DUP, INS, and INV were retained, with a minimum size threshold of 50 base pairs applied where applicable.

| Member | Technology | Caller | Raw records | Normalized count | SV type count |
|---|---|---|---|---|---|
| Patient | ONT | LongTR | 14352 | 18827 | BND=0; CSV=0; DEL=6250; DUP=0; INS=12577; INV=0 |
| | | SVIM-asm | 21740 | 18590 | BND=70; CSV=0; DEL=6754; DUP=31; INS=11700; INV=35 |
| | | cuteSV2 | 27773 | 27248 | BND=151; CSV=0; DEL=11790; DUP=135; INS=15120; INV=52 |
| | | Dysgu | 29452 | 27990 | BND=522; CSV=9; DEL=10766; DUP=177; INS=16372; INV=144 |
| | | svision | 27165 | 27137 | BND=0; CSV=96; DEL=11614; DUP=20; INS=15377; INV=30 |
| | SR | CNVnator | 20994 | 20994 | BND=0; CSV=0; DEL=17639; DUP=3355; INS=0; INV=0 |
| | | Manta | 175909 | 55588 | BND=43310; CSV=0; DEL=8376; DUP=1650; INS=2252; INV=0 |
| | OGM | Bionano | 5168 | 5166 | BND=322; CSV=0; DEL=1691; DUP=74; INS=3063; INV=16 |
| Mother | ONT | LongTR | 14530 | 19231 | BND=0; CSV=0; DEL=6402; DUP=0; INS=12829; INV=0 |
| | | SVIM-asm | 21340 | 18264 | BND=54; CSV=0; DEL=6661; DUP=19; INS=11499; INV=31 |
| | | cuteSV2 | 28588 | 28062 | BND=180; CSV=0; DEL=12150; DUP=167; INS=15509; INV=56 |
| | | Dysgu | 30050 | 28512 | BND=528; CSV=10; DEL=11069; DUP=161; INS=16601; INV=143 |
| | | SVision | 28530 | 28492 | BND=0; CSV=92; DEL=12273; DUP=20; INS=16069; INV=38 |
| | SR | CNVnator | 16408 | 16408 | BND=0; CSV=0; DEL=13599; DUP=2809; INS=0; INV=0 |
| | | Manta | 166470 | 54781 | BND=45346; CSV=0; DEL=6681; DUP=1267; INS=1487; INV=0 |
| | OGM | Bionano | 5244 | 5242 | BND=284; CSV=0; DEL=1754; DUP=69; INS=3120; INV=15 |
| Father | ONT | LongTR | 14367 | 18758 | BND=0; CSV=0; DEL=6191; DUP=0; INS=12567; INV=0 |
| | | SVIM-asm | 21482 | 18347 | BND=62; CSV=0; DEL=6696; DUP=22; INS=11539; INV=28 |
| | | cuteSV2 | 28630 | 27985 | BND=170; CSV=0; DEL=12130; DUP=167; INS=15457; INV=61 |
| | | Dysgu | 29516 | 27916 | BND=558; CSV=10; |

| | | | | | |
|---|---|---|---|---|---|
| | | | | | DEL=10801; DUP=176; INS=16247; INV=124 |
| | | SVision | 28765 | 28745 | BND=0; CSV=104; DEL=12341; DUP=19; INS=16246; INV=35 |
| | SR | CNVnator | 15608 | 15608 | BND=0; CSV=0; DEL=12933; DUP=2675; INS=0; INV=0 |
| | | Manta | 172070 | 64082 | BND=55464; CSV=0; DEL=6254; DUP=1119; INS=1245; INV=0 |
| | OGM | Bionano | 5151 | 5148 | BND=306; CSV=0; DEL=1678; DUP=79; INS=3069; INV=16 |

Supplementary Table 6. SVkhor merge and integration-stage counts for the clinical trio. Record counts are summarized following caller-level normalization, within-technology merging, sample-level integration of LR, SR, and OGM, and family-level cohort integration. LR and SR merged records represent the final svmerge clusters for each technology. OGM records are included directly after svnorm normalization. Sample-level integrated records represent the final techmerge output for each family member, while the family-level row indicates the final non-redundant catalog generated using SVkhor cohort-integration mode.

| SVkhor stage | Patient | Mother | Father | Total |
|---|---|---|---|---|
| Raw caller-level records | 322,553 | 311,160 | 315,589 | 949,302 |
| Normalized caller-level records | 201,540 | 198,992 | 206,589 | 607,121 |
| LR merged records | 58,674 | 60,381 | 60,197 | 179,252 |
| SR merged records | 73,202 | 69,100 | 77,646 | 219,948 |
| OGM normalized records | 5,166 | 5,242 | 5,148 | 15,556 |
| Sample-level integrated records | 128507 | 127,088 | 135,962 | 391,557 |
| Family-level cohort-integrated records | — | — | — | 317,857 |
